# Mortality associated with influenza and Omicron infections in France and vaccination of healthcare workers in nursing homes

**DOI:** 10.1101/2023.06.05.23290994

**Authors:** Edward Goldstein

**Affiliations:** Harvard Medical School, Boston, MA USA

## Abstract

**Background:** During the winter of 2022-2023, high rates of all-cause mortality, not seen since April 2020, were recorded in France, with excess all-cause mortality being related to the Omicron and influenza epidemics during that period. Moreover, that period saw a significant increase in the proportion of residents in long-term care facilities among cases of death in the population. Studies have found that increased influenza vaccination coverage in healthcare workers can result in a substantial reduction (up to 20%-30% during the course of select influenza seasons in the pre-pandemic period) in all-cause mortality in residents in nursing homes.

**Methods:** We applied the previously developed methodology to estimate the contribution of influenza infections to all-cause mortality in France for the 2014-2015 through the 2018-2019 influenza seasons, and the contribution of both SARS-CoV-2 and influenza infections to all-cause mortality between week 33, 2022 through week 12, 2023.

**Results:** For the 2014-2015 through the 2018-2019 seasons, influenza was associated with an average of 15654 (95% CI (13013,18340)) deaths, while between week 33, 2022 through week 12, 2023, we estimated 7851 (5213,10463) influenza-associated deaths and 32607 (20794,44496) SARS-CoV-2 associated deaths. The number of SARS-CoV-2-associated deaths during the Omicron epidemic was significantly higher than the number of deaths with COVID-19 listed on the death certificate or the hospitalization record – for example, between weeks 33-52 in 2022, we estimated 23983 (15307,32620) SARS-CoV-2-associated deaths in France, compared with 12811 deaths with COVID-19 listed on the death certificate, and 8639 in-hospital deaths with COVID-19 during the same period. Examination of US mortality data suggests a significant contribution of Omicron infections to mortality for cardiac disease and mental/behavioral disorders with COVID-19 not listed on the death certificate.

**Conclusions:** Our results suggest the need for boosting influenza vaccination coverage in different population groups (including healthcare workers, particularly nurse assistants for whom influenza vaccination coverage rates in France are low), as well as for wider use of influenza antiviral medications in influenza-related respiratory hospitalizations with different diagnoses (including pneumonia). Wider detection and treatment of Omicron infections, particularly in older individuals/persons with underlying health conditions such as cardiac disease and mental/behavioral disorders, and wider use of bivalent COVID-19 boosters would be needed in the event of the recrudescence of Omicron circulation in France.

## Introduction

While the risk of death for infections with the Omicron variant of SARS-CoV-2 in adults is significantly lower compared to the Delta variant [1,2], the appearance of the Omicron variant led to an increase in the proportion of severe outcomes (including hospitalizations and ICU admissions) in SARS-CoV-2-positive patients that were for a cause other than COVID-19 – see for example the data from France on hospitalizations and ICU admissions with a SARS-CoV-2 infection for a cause other than COVID-19 vs. admissions for COVID-19 [3]. This is related to differences in disease manifestation for Omicron infections vs. Delta infections for both emergency department (ED) admissions [4], hospitalizations [5], and admissions to critical care [6]. In particular, Omicron epidemics make a substantial contribution to mortality for cardiac causes, cancer, Alzheimer’s disease/neurological disorders, and other causes [7].

In December 2022, high rates of all-cause mortality, not seen since April 2020, were recorded in France [8]. Those high rates of excess all-cause mortality were related to the Omicron and the influenza epidemics during this period. However, levels of excess mortality observed in France during that period were only partly explained by the data on deaths with COVID-19 and influenza listed on death certificate (e.g. between weeks 33-52 in 2022, there were 12811 deaths with COVID-19 mentioned on the death certificate [9], and 554 deaths with influenza mentioned on the electronic death certificate [10]). For influenza, the fraction of associated deaths having influenza listed on the death certificate is lower than for Omicron – e.g. studies [11-13] suggest that only 23%-38% of influenza associated deaths have an underlying respiratory cause of death, for which only a further fraction has influenza listed on the death certificate, with an additional significant contribution of influenza infections to mortality for circulatory and other non-respiratory causes [11,14,15].

Data on the place of death in France suggest that the greatest relative increases in mortality in the 2^nd^ half of December 2022/1^st^ half of January 2023 took place in long-term care facilities for older individuals [16]. Epidemics of influenza in nursing homes can lead to high rates of incidence of infection [17], and vaccination of healthcare workers in nursing homes was shown to result in significant reduction in all-cause mortality in nursing home residents during select influenza seasons in the pre-pandemic period [18-20]. Influenza vaccination coverage in healthcare workers (HCWs) in France is relatively low [21,22], with particularly low vaccination coverage levels in nurse assistants (aide-soignant(e)) [21]. For the 2021-2022 season, influenza vaccination coverage in HCWs in residence establishments for dependent elderly persons (EHPAD) was estimated at 27.6% [22]. A study of COVID-19 in HCWs in France suggested that rates of infection in HCWs are by far the highest in nurse assistants [23]. It is likely that nurse assistants contribute significantly to the spread of influenza infections as well while having low vaccination coverage rates [21]. For Omicron infections, use of bivalent vaccines was associated with the reduction in the risk of death [24], while in nursing homes, in addition to vaccination of residents, vaccination of healthcare workers was also shown to reduce mortality rates in residents [25,26].

In addition to the benefits of increasing vaccination coverage in nursing home residents/staff, boosting vaccination coverage for both SARS-CoV-2 and influenza in other population groups should help mitigate the spread of those infections in the community and the associated burden of severe outcomes, including deaths. Vaccination coverage for the 2^nd^ booster for COVID-19 in France is largely restricted to persons aged over 60y, with relatively low coverage levels for boosters containing the Omicron components [27]. At the same time, there is evidence that vaccination reduces both the risk of acquisition of Omicron infections [28], as well as the risk of onward transmission of infection [29]. For influenza, vaccination coverage levels in different population groups in France are moderate-to-low, particularly in persons aged under 60y [30]. Studies have found that children experience higher than average influenza infection rates [31] and play an important role in the spread of influenza infection in different countries [32], including France [33]. Finally, rates of oseltamivir use for cases of severe influenza illness in France are moderate (e.g. 45.3% of cases in study [34]), and the use of oseltamivir in respiratory hospitalizations for other principal diagnoses (particularly pneumonia) that are accompanied by influenza infection can also reduce the risk of death [35].

In our earlier work [11,14], we developed a method for combing data on syndromic surveillance with data on virologic surveillance to estimate rates of mortality associated with the major influenza subtypes in the US. Subsequently, this method was applied to the estimation of influenza-associated mortality in other countries [36-38], including the EU population [38]. Here we adopt this framework to evaluate the contribution of influenza infections to all-cause mortality for the 2014-2015 through the 2018-2019 influenza seasons in France, as well as the contribution of SARS-CoV-2 and influenza infections to all-cause mortality in France between week 33, 2022 through week 12, 2023. The reason for omitting the period between 2020 through August 2022 from the analyses is that patterns of non-COVID-19 mortality have changed during the pandemic due to lockdowns, disruption in the circulation of other respiratory viruses [39] and other factors. For example, examination of mortality in England and Wales between March 2020 and June 2022 suggested that for deaths for which COVID-19 wasn’t coded as the underlying cause of death, there was a reduction of 36,7000 in the number of deaths for respiratory causes compared to averages during the previous 5 years [40]. Additionally, the heat wave in France resulted in significant levels of excess mortality in July 2022 [41]. However, starting mid-August 2023, baseline rates of mortality not associated with SARS-CoV-2 or influenza infections in France have returned to more regular patterns, which allowed for the inclusion of the period starting mid-August 2022 into the inference model developed in [11,14] to evaluate the contribution of influenza and SARS-CoV-2 infection to all-cause mortality in France. In addition to all-cause mortality data in France, we also examined data on US mortality with cardiovascular disease, as well as mortality with mental/behavioral disorders listed on the death certificate, with each of those data streams further stratified by having COVID-19 listed, or not listed on the death certificate [42]. The aim of that analysis was to examine how SARS-CoV-2 epidemic waves affected mortality for cardiac disease, and for mental/behavioral disorders, including mortality with COVID-19 not listed on the death certificate. The overall aim of this work is to help inform vaccination policies for SARS-CoV-2 and influenza, and policies for detecting/treating SARS-CoV-2 and influenza infections, particularly in persons with underlying health conditions.

## Methods

### Data availability

This manuscript is based on aggregate, de-identified publicly available data that can be accessed through refs. [8,9,42-45]. Data on weekly numbers of influenza-like illness (ILI) consultations in metropolitan France are available from the French sentinel surveillance [43]. Sentinel data on testing of respiratory specimens for the different influenza subtypes (A/H1N1, A/H3N2, B/Victoria and B/Yamagata) are available from WHO FluNET [44]. Data on the daily number of deaths in France starting 2015 are available from [8]. Data on population in France are available from [45]. Electronic records for deaths with COVID-19 listed on the death certificate are available from [9]. We note that many SARS-CoV-2-associated deaths are not listed in [9], and weekly death rates in [9] are used as a covariate in the regression model rather than the estimate of the contribution of COVID-19, or SARS-CoV2 infections to mortality. Finally, data on US mortality with different contributing causes are available from [42].

### Incidence indicators for the major influenza subtypes, and for SARS-CoV-2-associated deaths

Not all influenza-like illness (ILI) consultations in the sentinel data [43] correspond to influenza infections, and those that do, correspond to infection with different influenza subtypes. For each influenza subtype (e.g. A/H3N2), we define an indicator for the incidence of that subtype on week *t* (e.g. *A/H*3*N*2(*t*)) as

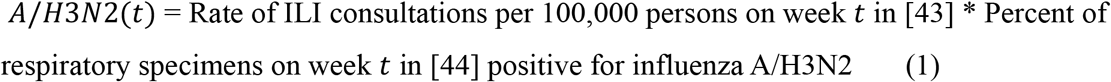

To relate the incidence indicators for the major influenza subtypes to weekly levels of all-cause mortality per 100,000 persons in France [8], we note that age distribution of influenza cases changes with the appearance of antigenically novel influenza strains, and this changes the relation between rates of influenza-associated ILI (incidence indicators in eq. 1) and rates of influenza-associated mortality. The relevant antigenic changes for our study period were (a) the appearance of an antigenically/genetically novel A/H3N2 strain during the 2014-2015 influenza season [46]; the appearance of the novel influenza B/Yamagata strain during the 2017-2018 season [38]; the 2018-2019 A/H3N2 epidemic strains belonging to several clades and exhibiting different immunity profiles for different birth cohorts [47]. Correspondingly, for the model relating A/H3N2 circulation to associated mortality, we split the A/H3N2 incidence indicator into three: *A/H*3*N*2_1_, equaling the A/H3N2 incidence indicator for the period from Jan. 2015 to Sep. 2015, and equaling to 0 for later weeks; *A/H*3*N*2_2_, equaling the A/H3N2 incidence indicator for the period from Oct. 2015 to Sep. 2018, and equaling to 0 for other weeks, and *A/H*3*N*2_3_, equaling the A/H3N2 incidence indicator for the period starting Oct. 2018, and equaling to 0 for other weeks. Similarly, we split the B/Yamagata incidence indicator into two, corresponding to the periods before and starting the 2017-2018 B/Yamagata epidemic. Finally, we note that it takes 1-2 weeks between influenza illness and influenza-associated mortality [11]. Correspondingly, we use a regression framework (eq. 2 below) to relate the mortality rate *M*(*t*) on week *t* to the *shifted* incidence indicators, e.g.

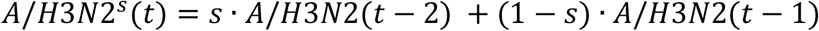

where the parameter *s* (common for all incidence indicators) is chosen to minimize the R-squared for the model fit. For the estimation of the rates of SARS-CoV-2-associated deaths, we use the weekly rates *Cov*_*e*_(*t*) of deaths with COVID-19 listed on the electronic death certificate in [9] as an indicator (covariate in the regression model).

### Regression model

The regression model that we use is:

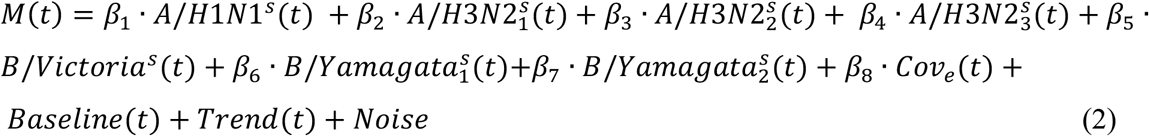

The time period that we include in the analysis is week 3, 2015 through week 2, 2020, and week 33, 2022 through week 12, 2023. The baseline represents weekly rates of mortality not associated with influenza or SARS-CoV-2 circulation, and *Baseline*(*t*) is modelled to have annual periodicity in week *t*. We use periodic cubic splines to model the baseline mortality rates whose shape is unknown [11,14]. The trend *Trend*(*t*) is modelled as a quadratic polynomial in week *t*. Finally, to account for the autocorrelation in the noise, we use a bootstrap procedure (resampling the noise on different weeks) to estimate the confidence bounds for various quantities evaluated in the model [11].

### US mortality

We use data on US mortality with cardiovascular disease (ICD-10 I00-I99), as well as mortality with mental/behavioral disorders (ICD-10 F01-F99) listed on the death certificate, with each of those data streams further stratified by having COVID-19 (ICD-10 U07.1) listed, or not listed on the death certificate [42].

## Results

### France mortality data

Figure 1 plots the results of the regression model in eq. 2. Figure 1 suggests that the model fits is generally temporally consistent, with baseline rates of non-influenza mortality + trend + the contribution of influenza infections to all-cause mortality (difference between the red and the green curves) explaining the patterns of mortality prior to the COVID-19 pandemic, while baseline + trend + contribution of influenza infections + contribution of SARS-CoV-2 infections to mortality explaining the patterns of mortality between week 33, 2022 and week 12, 2023 (with SARS-CoV-2 infections being responsible for most of excess mortality during that period).

**Figure 1:**
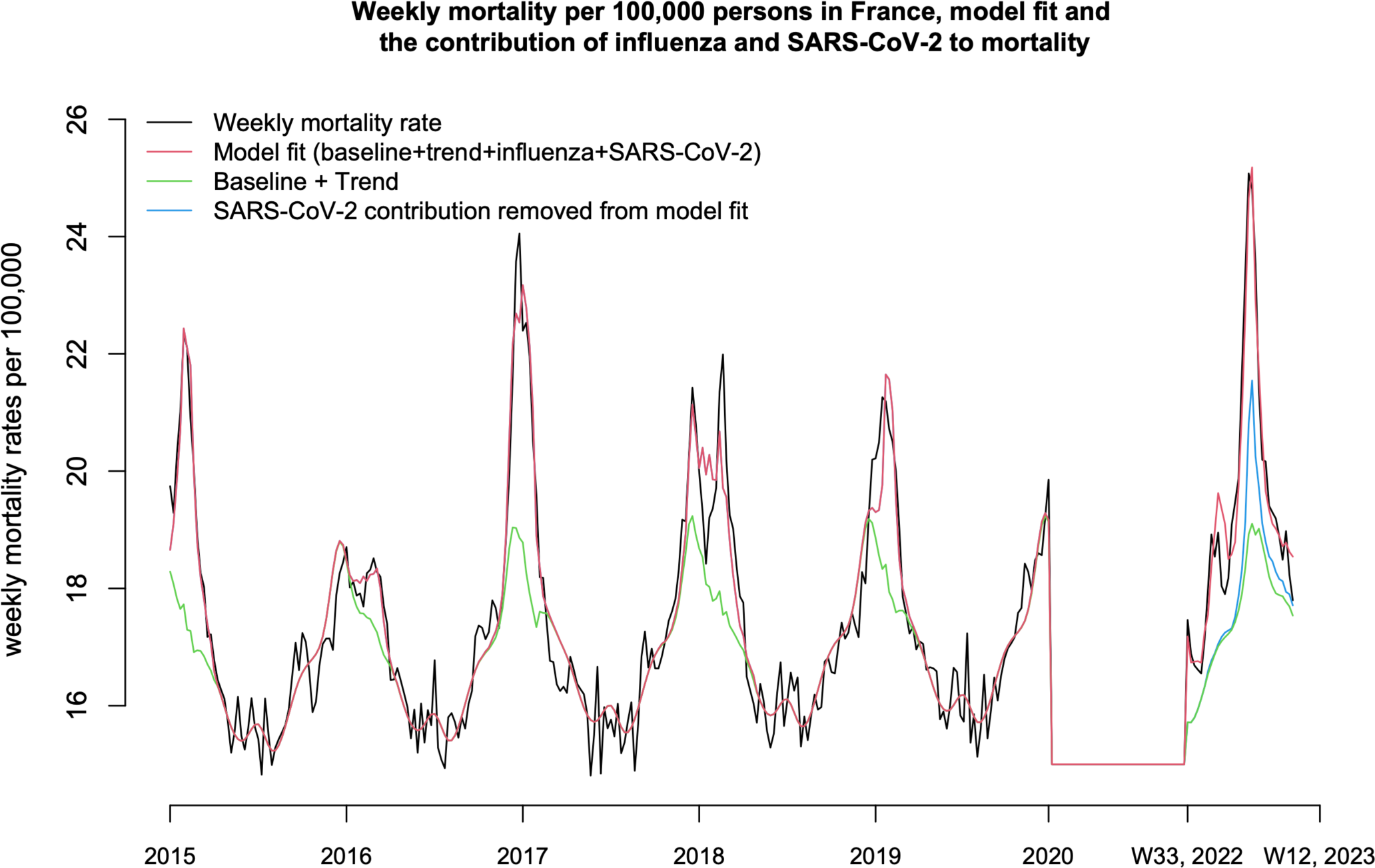
Weekly mortality rates for all causes between week 3, 2015 through week 2, 2020, and week 33, 2022 through week 12, 2023 (black curve), the baseline + trend for the rates of mortality not associated with influenza or SARS-CoV-2 infections in France (green curve), the model fit (baseline + trend + contribution of influenza infection +contribution of SARS-CoV-2 infection to mortality, red curve), and the model fit between week 33, 2022 through week 12, 2023 with the contribution of SARS-CoV-2 infection removed.

Table 1 gives the estimates of the contribution of influenza infections to all-cause mortality for the 2014-2015 through the 2018-2019 influenza seasons, and the contribution of SARS-CoV-2 and influenza infections to all-cause mortality between week 33, 2022 through week 12, 2023. For the 2014-2015 through the 2018-2019 seasons, influenza was associated with an average of 15654 (95% CI (13013,18340)) deaths, while between week 33, 2022 through week 12, 2023, we estimated 7851 (5213,10463) influenza-associated deaths and 32607 (20794,44496) SARS-CoV-2 associated deaths.

**Table 1:** The contribution of influenza infection to all-cause mortality for the 2014-2015 through the 2018-2019 influenza seasons, and the contribution of SARS-CoV-2 and influenza infections to all-cause mortality between week 33, 2022 through week 12, 2023.

| Season | Influenza-associated deaths | SARS-CoV-2-associated deaths |
| --- | --- | --- |
| 2014-2015 | 19779 (15438,24122) |  |
| 2015-2016 | 5432 (902,9932) |  |
| 2016-2017 | 21997 (17891,26067) |  |
| 2017-2018 | 19138 (13599,24838) |  |
| 2018-2019 | 11925 (8632,15245) |  |
| Annual average, 2014-2015 through 2018-2019 seasons | 15654 (13013,18340) |  |
| Week 33, 2022 – Week 12, 2023 | 7851 (5213,10463) | 32607 (20794,44496) |

Data on mortality with COVID-19 listed on the death certificate in France are available through the end of 2022 [9]. For the period between weeks 33-52 in 2022, we estimated 23983 (15307,32620) SARS-CoV-2-associated deaths in France, compared with 12811 deaths with COVID-19 listed on the death certificate [9], and 8639 in-hospital deaths with COVID-19 during the same period [48].

### US mortality data

Figure 2 plots the weekly number of US deaths with a cardiac cause but without COVID-19 listed on death certificate, as well as the number of deaths with both a cardiac cause and COVID-19 listed on the death certificate between Nov. 21, 2020—Nov. 19, 2022. Figure 3 plots the weekly number of US deaths with mental/behavioral disorders but without COVID-19 listed on death certificate, as well as the number of deaths with both mental/behavioral disorders and COVID-19 listed on the death certificate between Nov. 21, 2020—Nov. 19, 2022. Figures 2 and 3 suggest that compared to the SAR-CoV-2 epidemic in the Fall-Winter of 2021-2022, for the first Omicron epidemic wave in 2022 there were significantly higher increases in mortality with cardiovascular disease, as well as in mortality with mental/behavioral disorders listed on the death certificate and COVID-19 NOT listed on the death certificate, and smaller increases (for the first Omicron epidemic wave compared to the 2020-2021 epidemic) in mortality with either cardiovascular disease and COVID-19 listed on the death certificate, or with mental/behavioral disorders and COVID-19 listed on the death certificate.

**Figure 2:**
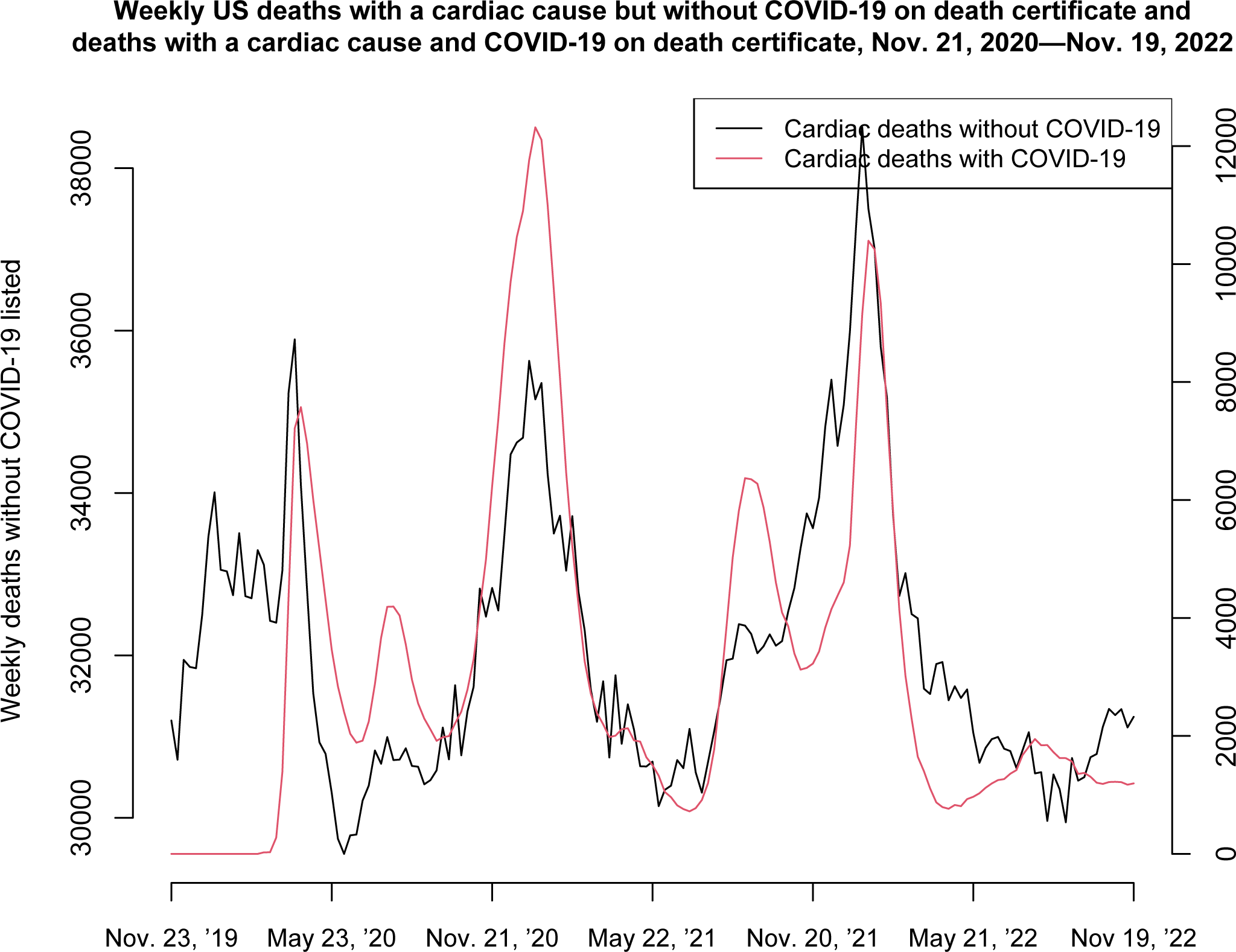
Weekly US deaths with a cardiac cause but without COVID-19 on death certificate and deaths with a cardiac cause and COVID-19 on death certificate, Nov. 21, 2020—Nov. 19, 2022

**Figure 3:**
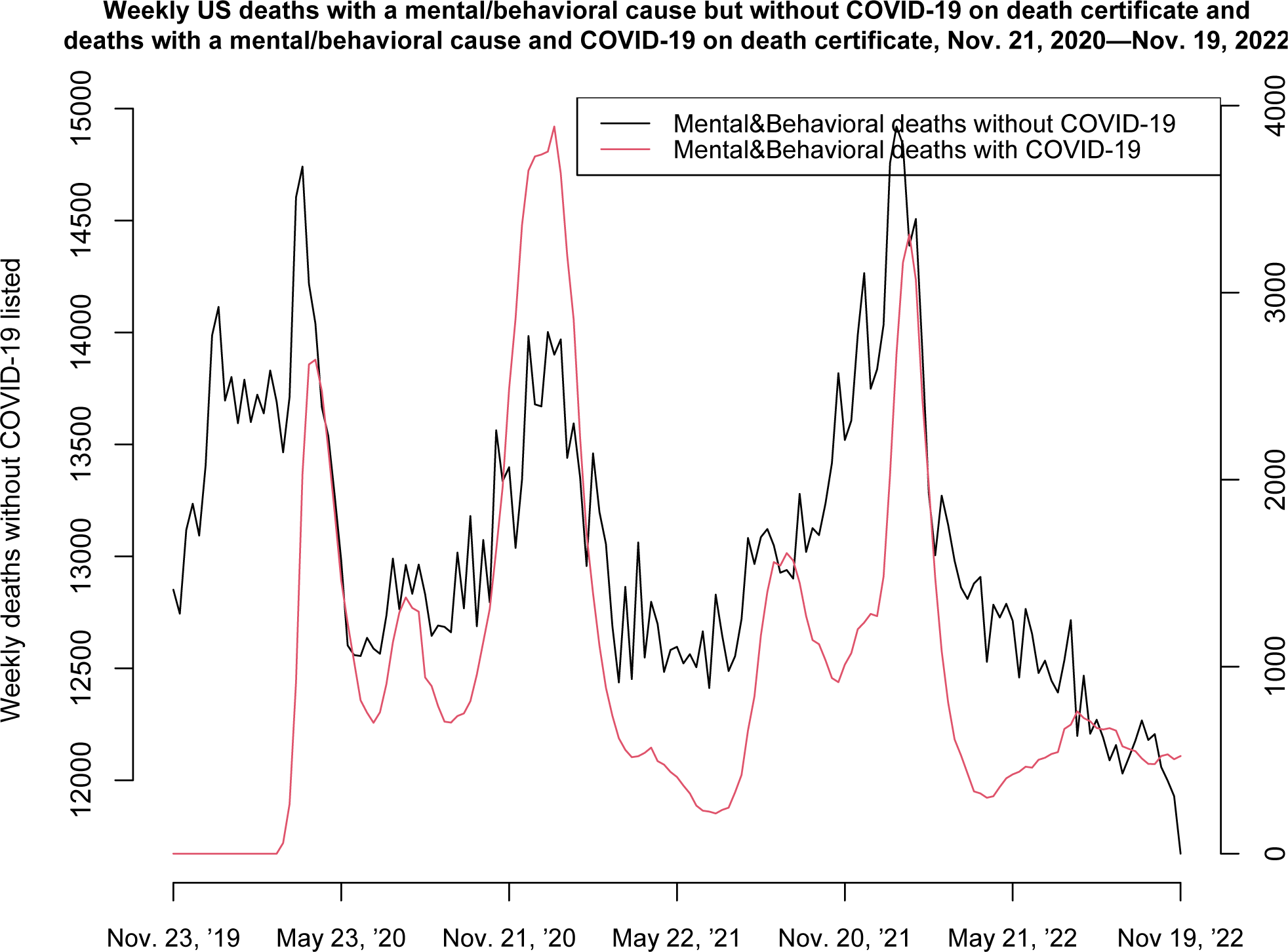
Weekly US deaths with mental/behavioral disorders but without COVID-19 on death certificate and deaths with mental/behavioral disorders and COVID-19 on death certificate, Nov. 21, 2020—Nov. 19, 2022

## Discussion

During the winter of 2022-2023, high levels of excess mortality (not seen since the 1^st^ COVID-19 wave in April 2020) were recorded in France [8], with excess mortality being primarily associated with the Omicron epidemic [3], but also the influenza epidemic during that period [49]. Additionally, the greatest increases in mortality during that period were recorded in residents in long-term care facilities [16]. Here, we applied the previously developed regression model to French mortality data between 2015-2019, as well as data between week 33, 2022 -- week 12, 2023 to estimate the contribution of both influenza and Omicron infections to all-cause mortality in France. We found that for the 2014-2015 through the 2018-2019 seasons, influenza was associated with an average of 15654 (95% CI (13013,18340)) deaths (with an additional 7851 (5213,10463) influenza-associated deaths during the 2022-2023 season, with short-term cross-immunity from Omicron infections potentially mitigating the influenza epidemic somewhat during that period [50]). We also estimated 32607 (20794,44496) SARS-CoV-2 associated deaths between week 33, 2022 – week 12, 2023. The number of SARS-CoV-2-associated deaths during the Omicron epidemic was significantly higher than the number of deaths with COVID-19 listed on the death certificate or the hospitalization record – for example, between weeks 33-52 in 2022, we estimated 23983 (15307,32620) SARS-CoV-2-associated deaths in France, compared with 12811 deaths with COVID-19 listed on the death certificate, and 8639 in-hospital deaths with COVID-19 during the same period.

During the pre-pandemic period, residents in establishments for dependent elderly persons in France (EHPAD) made up about a 1/4 of all deaths in France, e.g. [51,52]. During influenza and Omicron epidemics, the relative contribution of EHPAD to mortality increases, as evidenced by the data during winter 2022-2023. Influenza outbreaks in nursing homes tend to have higher attack rate compared to infection rates in the overall population of elderly individuals. For example, study [17] suggests that for the 49 outbreaks caused by influenza, the median attack rate in residents was 33% (range 4-94%), with a median case-fatality rate for residents being 6.5%. A meta-analysis that included 4 cluster randomized trials and 4 observational studies on influenza vaccination in HCWs conducted in long-term care or hospital settings found that pooled risk ratios across trials for all-cause mortality and influenza-like illness in patients/residents were 0.71 (95% confidence interval [CI], .59-.85) and 0.58 (95% CI, .46-.73), respectively; pooled estimates for laboratory-confirmed influenza were not statistically significant [18]. Study [19] found that vaccination of healthcare workers (HCWs) was associated with reduction in all-cause mortality in residents/patients (VE 40%, 95% CI 27% to 50%) and deaths from pneumonia (VE 39%, 95% CI 2% to 62%). A more recent cluster-randomized trial of influenza vaccination in nursing home staff showed a 20% lower mortality in residents (P=.02) [20]. While magnitudes of the above estimates of the effect of HCW vaccination on all-cause mortality in residents in long-term care facilities are quite high, the effect varies year-to-year (as suggested e.g. by the results for two consecutive influenza seasons in the study [53]). The findings in studies [18-20] support the case for increasing influenza vaccination rates in HCWs, particularly nurse assistants [21,22]. Finally, some studies suggest limited effect of influenza vaccination in HCWs on lab-confirmed influenza illness in residents [17,54]. However, lab-confirmed influenza illness is *not* an objective outcome as most episodes of influenza infection are not being tested for/detected. In the studies that examine the effect of influenza vaccination in healthcare workers [18-20], some of the facilities were randomized to receive influenza vaccination in HCWs, while others did not receive a placebo; it is not unlikely that facilities randomized for HCW vaccination also started to test for influenza infection more frequently as a result of being selected for HCW vaccination, whereas facilities that weren’t selected for HCW vaccination might have tested their residents less frequently so not to exhibit a high burden of influenza infection compared to facilities that were selected for vaccination in those trials. Thus, estimates of effect of HCW vaccination on lab-confirmed influenza illness are difficult to interpret, with vaccination of HCWs found to be associated with a significant reduction in ILI in nursing home residents in study [20].

Our finding about the higher number of SARS-CoV-2-associated deaths in France compared to the number of deaths with COVID-19 listed on the death certificate (1^st^ paragraph of Discussion) is in agreement with a significant contribution of Omicron infections to mortality for cardiac causes, cancer, Alzheimer’s disease/neurological disorders and other causes found in [7]. Data on US mortality examined in this paper also suggests a significant burden of excess deaths for cardiac disease and for mental/behavioral disorders without COVID-19 listed on the death certificate during the Omicron epidemic wave in 2022 (Figure 2,3). Those findings suggest the need to widen the detection/treatment of Omicrons infections, particularly in individuals with underlying health conditions, as well as the need to increase COVID-19 booster vaccination coverage in the whole population to mitigate SARS-CoV-2 transmission in the community. Our findings about the relatively high rates of influenza-associated mortality in France (e.g. compared to the estimates for the EU [38]) support the need for increasing influenza vaccination coverage in different population groups besides healthcare workers with the aim of mitigating influenza transmission in the community, particularly since influenza vaccine effectiveness in older individuals can be quite low [55]. Influenza vaccination coverage in non-elderly individuals in France is significantly lower compared to the US [30,56], with children known to play a significant role in influenza transmission in the community [31-33]. Finally, detection and treatment of influenza infection in hospitalizations for respiratory causes, including pneumonia, was found to have benefits in terms of reducing mortality rates in hospitalized patients [35], with a major increase in the proportion of pneumonia patients who were tested for influenza infection in the US taken place in recent years [35].

Our results have some limitations. Influenza surveillance data in France pertain to mainland France [42], whereas we’ve used data on mortality for the whole of France. Additionally, sentinel data on testing for viral specimens [43] has a moderate sample size and may not represent all of France. We note that influenza epidemics exhibit a great deal of temporal synchrony [57,58] which should help address the above limitations. Finally, despite the fact that we split some of the influenza subtype incidence indicators into several time periods, where might still be temporal variability in the relation between the incidence indicators used in this paper and rates of associated mortality. For example, while model fits are generally temporally consistent (Figure 1), the model fit for the mortality data for the 2017-2018 season is worse compared to other influenza seasons, which might be related to the fact that the influenza subtypes that circulated during that season (A/H1N1 and B/Yamagata) have different age distributions feeding into one ILI data stream.

## Conclusions

Our findings about influenza-associated mortality in France, as well as the low influenza vaccination rates in healthcare workers in France [21,22] suggest the need for boosting those vaccination rates. In the event of the recrudescence of Omicron circulation in France, efforts should be undertaken to administer booster vaccination for COVID-19 to HCWs, with additional benefits being offered by bivalent booster vaccines [24] and with coverage levels for booster vaccines containing the Omicron component in healthcare workers in France being relatively low [3]. Wider detection and treatment of Omicron infections, particularly in older individuals/persons with underlying health conditions such as cardiac disease and mental/behavioral disorders [7] should help mitigate the mortality burden of future Omicron epidemic waves. Finally, our results suggest that boosting influenza vaccination coverage in different population groups (including children [31-33]) and wider testing for influenza infection in respiratory illness episodes with different principal diagnoses (including pneumonia) during periods of active influenza circulation in combination with the use of antiviral medications [35] should be considered with the aim of mitigating the mortality burden of future influenza epidemics.

## Data Availability

This manuscript is based on aggregate, de-identified publicly available data that can be accessed through refs. [8,9,42-45].

https://wonder.cdc.gov/mcd-icd10-provisional.html

https://www.sentiweb.fr/france/fr/?page=table

https://app.powerbi.com/view?r=eyJrIjoiNjViM2Y4NjktMjJmMC00Y2NjLWFmOWQtODQ0NjZkNWM1YzNmIiwidCI6ImY2MTBjMGI3LWJkMjQtNGIzOS04MTBiLTNkYzI4MGFmYjU5MCIsImMiOjh9

https://www.insee.fr/fr/statistiques?debut=0&theme=1

https://opendata.idf.inserm.fr/cepidc/covid-19/telechargements

## References

[1] Nyberg T, et al. Comparative analysis of the risks of hospitalization and death associated with SARS-CoV-2 omicron (B.1.1.529) and delta (B.1.617.2) variants in England: a cohort study. The Lancet 2022; 399(10332): 1303–1312

[2] Auvigne V, et al. Severe hospital events following symptomatic infection with Sars-CoV-2 Omicron and Delta variants in France, December 2021-January 2022: A retrospective, population-based, matched cohort study. EClinicalMedicine. 2022;48:101455

[3] Santé publique France. Santé publique France. InfoCovidFrance: Chiffres clés et évolution de la COVID-19 en France et dans le Monde. 2022. Available from: https://www.santepubliquefrance.fr/dossiers/coronavirus-covid-19/coronavirus-chiffres-cles-et-evolution-de-la-covid-19-en-france-et-dans-le-monde

[4] Bouzid D, et al. Comparison of Patients Infected With Delta Versus Omicron COVID-19 Variants Presenting to Paris Emergency Departments: A Retrospective Cohort Study. Ann Intern Med. 2022;175(6):831–837

[5] Leiner J, et al. Characteristics and outcomes of COVID-19 patients during B.1.1.529 (Omicron) dominance compared to B.1.617.2 (Delta) in 89 German hospitals. BMC Infect Dis. 2022;22(1):802

[6] de Prost N, et al. Clinical phenotypes and outcomes associated with SARS-CoV-2 variant Omicron in critically ill French patients with COVID-19. Nat Commun. 2022;13:6025.

[7] Australian Bureau of Statistics. COVID-19 Mortality by wave. 2023. Available from: https://www.abs.gov.au/articles/covid-19-mortality-wave

[8] Institut national de la statistique et des études économiques (INSEE). Nombre de décès quotidiens France, régions et départements. 2023. https://www.insee.fr/fr/statistiques/4931039?sommaire=4487854#tableau-figure1

[9] The Institut national de la santé et de la recherche médicale (INSERM). Données sur la Covid-19 du centre d’épidémiologie sur les causes médicales de décès de l’Inserm. 2022. Available from: https://opendata.idf.inserm.fr/cepidc/covid-19/telechargements

[10] Santé publique France. Bulletin épidémiologique grippe, semaine 52. Saison 2022-2023. 2023. https://www.santepubliquefrance.fr/maladies-et-traumatismes/maladies-et-infections-respiratoires/grippe/documents/bulletin-national/bulletin-epidemiologique-grippe-semaine-52.-saison-2022-2023

[11] Goldstein E, Viboud C, Charu V, et al. Improving the estimation of influenza-related mortality over a seasonal baseline. Epidemiology. 2012;23(6):829–38.

[12] Hansen CL, Chaves SS, Demont C, Viboud C. Mortality associated with influenza and respiratory syncytial virus in the US, 1999-2018. JAMA Netw Open. 2022;5(2):e220527

[13] Schmidt SSS, Iuliano AD, Vestergaard LS, Mazagatos-Ateca IC, Larrauri A, M. Brauner JM, et al. All-cause versus cause-specific excess deaths for estimating influenza-associated mortality in Denmark, Spain, and the United States. Influenza Other Respir Viruses. 2022 Jul; 16(4): 707–716

[14] Quandelacy TM, Viboud C, Charu V, et al. Age- and Sex-related Risk Factors for Influenza-associated Mortality in the United States Between 1997-2007. Am J Epidemiol. 2014;179(2):156–67

[15] Kwong JC, Schwartz KL, Campitelli MA, et al. Acute Myocardial Infarction after Laboratory-Confirmed Influenza Infection. N Engl J Med. 2018;378(4):345–353

[16] Institut national de la statistique et des études économiques (INSEE). Daily deaths files broken down by sex, age and place of death. https://www.insee.fr/en/statistiques/4493808?sommaire=4493845

[17] Lansbury LE, Brown CS, Nguyen-Van-Tam JS. Influenza in long-term care facilities. Influenza Other Respir Viruses. 2017;11(5):356–366

[18] Ahmed F, Lindley MC, Allred N, Weinbaum CM, Grohskopf L. Effect of influenza vaccination of healthcare personnel on morbidity and mortality among patients: systematic review and grading of evidence. Clin Infect Dis. 2014;58(1):50–7

[19] Thomas RE, Jefferson T, Demicheli V, Rivetti V. Influenza vaccination for healthcare workers who work with the elderly. Cochrane Database Syst Rev. 2006;(3):CD005187

[20] Lemaitre M, Meret T, Rothan-Tondeur M, Belmin J, Lejonc JL, Luquel L, et al. Effect of influenza vaccination of nursing home staff on mortality of residents: a cluster-randomized trial. J Am Geriatr Soc. 2009;57(9):1580–6

[21] Santé Publique France. Couverture vaccinale antigrippale chez les professionnels de santé. October 2019. https://www.santepubliquefrance.fr/determinants-de-sante/vaccination/documents/bulletin-national/bulletin-de-sante-publique-vaccination.-octobre-2019

[22] Santé Publique France. Quelle est la couverture vaccinale contre la grippe des résidents et des professionnels en établissements médico-sociaux ? Point au 1er juin 2022. https://www.santepubliquefrance.fr/maladies-et-traumatismes/maladies-et-infections-respiratoires/grippe/documents/enquetes-etudes/quelle-est-la-couverture-vaccinale-contre-la-grippe-des-residents-et-des-professionnels-en-etablissements-medico-sociaux-point-au-1er-juin-2022

[23] Belan M, Charmet T, Schaeffer L, Tubiana S, Duval X, Lucet J-C, et al. SARS-CoV-2 exposures of healthcare workers from primary care, long-term care facilities and hospitals: a nationwide matched case-control study. Clin Microbiol Infect. 2022 Nov;28(11):1471–1476

[24] Johnson AG, Linde L, Ali AR, DeSantis A, Shi M, Adam C, et al. COVID-19 Incidence and Mortality Among Unvaccinated and Vaccinated Persons Aged ≥12 Years by Receipt of Bivalent Booster Doses and Time Since Vaccination — 24 U.S. Jurisdictions, October 3, 2021–December 24, 2022. MMWR Morb Mortal Wkly Rep. 2023;72(6):145–152

[25] Sinha S, Konetzka RT. Association of COVID-19 Vaccination Rates of Staff and COVID-19 Illness and Death Among Residents and Staff in US Nursing Homes. JAMA Netw Open. 2022 Dec 1;5(12):e2249002

[26] McGarry BE, Barnett ML, Grabowski DC, Gandhi AD. Nursing Home Staff Vaccination and Covid-19 Outcomes. N Engl J Med. 2022;386(4):397–398

[27] Gouvernement de la République française. Vaccins (COVID-19). 2022. Available from: https://www.gouvernement.fr/info-coronavirus/vaccins

[28] Chin ET, Leidner D, Lamson L, Lucas K, Studdert DM, Goldhaber-Fiebert JD, et al. Protection against Omicron from Vaccination and Previous Infection in a Prison System. N Engl J Med. 2022 Nov 10;387(19):1770–1782

[29] Tan ST, Kwan AT, Rodríguez-Barraquer I, Singer BJ, Park HJ, Lewnard JA, et al. Infectiousness of SARS-CoV-2 breakthrough infections and reinfections during the Omicron wave. Nature Medicine 2023;29:358–365

[30] Robert J, Detournay B, Levant MC, Uhart M, Gourmelen J, Cohen JM. Flu vaccine coverage for recommended populations in France. Med Mal Infect. 2020;50(8):670–675

[31] Monto A, Koopman J, Longini IM., Jr Tecumseh study of illness. XIII. Influenza infection and disease, 1976–1981. Am J Epidemiol. 1985;102:553–563

[32] Worby CJ, Chaves SS, Wallinga J, Lipsitch M, Finelli L, Goldstein E. On the relative role of different age groups in influenza epidemics. Epidemics 2015;13:10–16.

[33] Cauchemez S, Valleron AJ, Böelle PY, Flahaut A, Ferguson NM. Estimating the impact of sc3ool closure on influenza transmission from Sentinel data. Nature. 2008;452:750–754.

[34] Cizeron A, Saunier F, Gagneux-Brunon A, Pillet S, Cantais A, Botelho-Nevers E. Low rate of oseltamivir prescription among adults and children with confirmed influenza illness in France during the 2018-19 influenza season. J Antimicrob Chemother. 2021;76(4):1057–1062

[35] Deshpande A, Klompas M, Yu P-C, Imrey PB, Pallotta AM, Higgins T, et al. Influenza Testing and Treatment Among Patients Hospitalized With Community-Acquired Pneumonia. Chest. 2022;162(3):543–555

[36] Pebody RG, Green HK, Warburton F, et al. Significant spike in excess mortality in England in winter 2014/15 - influenza the likely culprit. Epidemiol Infect. 2018;146(9):1106–1113

[37] Rosano A, Bella A, Gesualdo F, et al. Investigating the impact of influenza on excess mortality in all ages in Italy during recent seasons (2013/14-2016/17 seasons). Int J Infect Dis. 2019;88:127–134

[38] Nielsen J, Vestergaard LS, Richter L, Schmid D, Bustos N, Asikainen T, et al. European all-cause excess and influenza-attributable mortality in the 2017/18 season: should the burden of influenza B be reconsidered? Clin Microbiol Infect. 2019 Oct;25(10):1266–1276.

[39] Editorial. COVID-19 pandemic disturbs respiratory virus dynamics. The Lancet Respiratory Medicine. 2022;10(8)725

[40] Office for National Statistics, UK. Excess deaths in England and Wales: March 2020 to June 2022. 20202. Available from: https://www.santepubliquefrance.fr/dossiers/coronavirus-covid-19/coronavirus-chiffres-cles-et-evolution-de-la-covid-19-en-france-et-dans-le-monde

[41] Santé Publique France. Bulletin de santé publique canicule. Bilan été 2022. Available from: https://www.santepubliquefrance.fr/determinants-de-sante/climat/fortes-chaleurs-canicule/documents/bulletin-national/bulletin-de-sante-publique-canicule.-bilan-ete-2022

[42] US Centers for Disease Control and Prevention. CDC Wonder mortality data. Available from: https://wonder.cdc.gov/mcd-icd10-provisional.html

[43] Réseau Sentinelles. France. Surveillance continue n.d. 2022. https://www.sentiweb.fr/france/fr/?page=table (Accessed on Nov. 2022)

[44] World Health Organization. Influenza Virus Detections reported to FluNET. 2022. https://app.powerbi.com/view?r=eyJrIjoiNjViM2Y4NjktMjJmMC00Y2NjLWFmOWQtODQ0NjZkNWM1YzNmIiwidCI6ImY2MTBjMGI3LWJkMjQtNGIzOS04MTBiLTNkYzI4MGFmYjU5MCIsImMiOjh9

[45] Institut national de la statistique et des études économiques (INSEE). Évolution et structure de la population. 2022. Available from: https://www.insee.fr/fr/statistiques?debut=0&theme=1

[46] Skowronski DM, et al. (2016) A Perfect Storm: Impact of Genomic Variation and Serial Vaccination on Low Influenza Vaccine Effectiveness During the 2014–2015 Season. Clin Infect Dis.;63(1):21–32

[47] Kissling E, Pozo F, Buda S, Vilcu A, Gherasim A, Brytting M, et al. Low 2018/19 vaccine effectiveness against influenza A(H3N2) among 15–64-year-olds in Europe: exploration by birth cohort. Euro Surveill. 2019;24(48).

[48] Santé Publique France. Données hospitalières relatives à l’épidémie de COVID-19. Available from: https://www.data.gouv.fr/fr/datasets/donnees-hospitalieres-relatives-a-lepidemie-de-covid-19/

[49] Santé Publique France. Bulletin épidémiologique grippe, semaine 15. Saison 2022-2023. 2023. https://www.santepubliquefrance.fr/maladies-et-traumatismes/maladies-et-infections-respiratoires/grippe/documents/bulletin-national/bulletin-epidemiologique-grippe-semaine-15.-saison-2022-2023

[50] Deleveaux S, Clarke-Kregor A, Fonseca-Fuentes X, Mekhaiel E. Exploring the Possible Phenomenon of Viral Interference Between the Novel Coronavirus and Common Respiratory Viruses. J Patient Cent Res Rev. 2023;10(2):91–97

[51] Direction de la recherche, des études, de l’évaluation et des statistiques (DREES). L’Ehpad, dernier lieu de vie pour un quart des personnes décédées en France en 2015. 2018. https://drees.solidarites-sante.gouv.fr/sites/default/files/er1094_toile.pdf

[52] Botton J, Drouin J, Bertrand M, Jabagi M-J, Weill A, Zureik M, et al. Fréquence des décès et des hospitalisations chez les résidents des établissements d’hébergement pour personnes âgées dépendantes (EHPAD) et des unités de soin longue durée (USLD) en France au cours des années 2018 et 2019. EPI-PHARE - Groupement d’intérêt scientifique (GIS). 2021. https://www.epi-phare.fr/app/uploads/2021/02/epi-phare_rapport_deces_hospit_hors-ehpad_20210205.pdf

[53] Hayward AC, Harling R, Wetten S, Johnson AM, Munro S, Smedley J, et al. Effectiveness of an influenza vaccine programme for care home staff to prevent death, morbidity, and health service use among residents: cluster randomised controlled trial. BMJ. 2006 Dec 16;333(7581):1241

[54] Thomas RE, Jefferson T, Lasserson TJ. Influenza vaccination for healthcare workers who care for people aged 60 or older living in long-term care institutions. Cochrane Database Syst Rev. 2016;2016(6):CD005187

[55] US Centers for Disease Control and Prevention. Past Seasons Vaccine Effectiveness Estimates. Available from: https://www.cdc.gov/flu/vaccines-work/past-seasons-estimates.html

[56] US Centers for Disease Control and Prevention. Flu Vaccination Coverage, United States, 2019–20 Influenza Season. https://www.cdc.gov/flu/fluvaxview/coverage-1920estimates.htm

[57] Schanzer DL, Langley JM, Dummer T, et al. The Geographic Synchrony of Seasonal Influenza A Waves across Canada and the United States. PLoS One 2011; 6(6): e21471

[58] Morris SE, Freiesleben de Blasio D, Viboud C, et al. Analysis of multi-level spatial data reveals strong synchrony in seasonal influenza epidemics across Norway, Sweden, and Denmark. PLoS One. 2018 May;13(5):e0197519

